# Preparing for Disease X requires CLARITY – Lessons learned from COVID-19 – A systematic Review

**DOI:** 10.1101/2024.11.27.24317926

**Authors:** Michael Boivin, Jean Bourbeau, Maria Sedeno, Emily Horvat, Pei-Zhi Li, Baraa Noueihed, Kim A. Connelly, Akshay B. Jain, Peter Lin, Bruce D. Mazer, Gustavo Saposnik, William D. Strain

## Abstract

**Objectives:** The COVID-19 pandemic has highlighted the critical need for comprehensive preparedness strategies for future pandemics, particularly those caused by unknown pathogens, or Disease X.

**Study Design:** Our systematic review examined current literature on COVID-19 risk factors, identifying significant gaps and inconsistencies in data reporting.

**Methods:** A systematic literature search was conducted including all English language published retrospective and prospective observational studies which documented clinical outcomes of COVID-19 in adult patients, on Ovid MEDLINE(R) and Epub databases (December 2019 to March 2023).

**Results:** The search yielded 440 articles of which 29 were included in the systematic review. We identified major risk factors for severe outcomes. However, inconsistencies in reporting comorbidities, age, disease control levels, medication use, vaccination status, and variant type, limited our analysis. In many publications, this information was not included. Additionally, variations in public attitudes, knowledge, and behaviours regarding COVID-19 vaccines further complicated resource distribution. We propose the CLARITY model to address these gaps by standardizing risk factor reporting, encompassing detailed comorbidity profiles, disease control metrics, age analyses, medication and vaccination data, and variant-specific information.

**Conclusions:** Implementing the CLARITY model could improve preparedness and response strategies for Disease X, enabling healthcare professionals and systems to allocate resources more effectively and mitigate the impact of future pandemics. This review underscores the necessity for a coordinated global effort to enhance pandemic preparedness through improved data reporting and risk stratification.

**Funding:** This study received financial support from Moderna Biopharma Canada Corporation.

**Trial Registration:** PROSPERO Identifier CRD42023449647

## 1. Introduction

The COVID-19 pandemic and the spread of SARS-CoV-2 virus has demonstrated human vulnerability to emerging diseases. Future pandemics are highly probable as there are an estimated 1.67 million unknown viruses which exist in mammals and birds [1]. Estimates are that up to 827,000 of these viruses are likely to infect people [1]. To adequately prepare for the next pandemic, the World Health Organization implemented a Research and Development Blueprint to decrease the time for development, assessment and countermeasures for the world’s most dangerous pathogens [2]. The goal was to develop a preparedness for Disease X. This condition is caused by Pathogen X, an infectious agent that is currently not known to cause human disease, but could be associated with a future epidemic or pandemic [2].

Although identification of Pathogen X is important, the COVID-19 pandemic demonstrated that healthcare systems and resources could be quickly overwhelmed as the infection infects an increasing number of the population [3]. Healthcare providers, systems and governments experienced shortages in resources (e.g. hospital beds, ventilators, masks, therapeutics) that significantly hampered pandemic mitigation strategies and the management of people infected with the virus [3]. This stressed the importance of the utilization of these limited resources on the individuals who were at highest risk of severe outcomes and those who could benefit the most from these interventions. Although risk factors for severe COVID-19 were quickly identified at the start of the pandemic, there was a lack of clarity of the extent of these risk factors on the risk of severe outcomes. For example, many conditions were associated with a higher risk of severe COVID-19, but additional factors such as the patient’s additional comorbidities, current therapy, level of disease control and vaccination status were commonly not reported in much of the COVID-19 risk factor literature.

To provide additional clarity in the risk for severe COVID-19 and gaps in the literature, our group undertook a systematic review and meta-analysis to identify individuals at high risk of adverse outcomes from COVID-19. We also propose the use of a standardized reporting process for evaluating risk factors for future pandemics to provide researchers, healthcare professionals, healthcare systems and governments with further clarity on how to allocate resources to manage a future pandemic caused by Pathogen X.

## 2. Materials and Methods

A systematic literature search was conducted (PROSPERO Identifier CRD42023449647) to identify manuscripts specific to COVID-19 and high-risk populations. The supplementary materials include additional Information on Methodology, a detailed account of the literature search strategy (**Table S3**), PRISMA Checklists (**Tables S1** and **S2**), study eligibility and selection criteria, data extraction procedures, and statistical analysis methods.

## 3. Results

The search of Ovid MEDLINE(R) and Epub databases yielded 440 articles documenting clinical outcomes of COVID-19 in relation to comorbidities. Following the initial abstract screening, 52 studies were eligible for full-text screening assessment. A total of 23 studies were excluded because the reported outcomes (n=8) or study design (n=12) did not meet our inclusion criteria, were missing information that prohibited data extraction and analysis (n=2) or included pediatric population higher than 10% representation (n=1).

Twenty-nine (29) full-text studies were used for data extraction, of which, 3 studies reported mortality rates for COVID-19 positive cases [4–6], 21 studies documented hospitalization rates including intensive care unit (ICU) admission, intubation, and oxygen therapy [7–27], and 5 studies reported both [28–32]. The PRISMA flowchart for study selection and additional characteristics of included studies are shown in **Figure S1** and **Table S4** of the Supplement.

### 3.1. Risk of hospitalization

Twenty-six studies reported hospitalization rates which includes ICU admission, intubation, and/or non-invasive ventilation support [7–32]. **Table 1** presents the demographics of hospitalized and non-hospitalized patients for studies reporting mean values. A higher proportion of hospitalized patients were male, older, and tended to be active smokers. COVID-19 positive patients aged ≥ 65 years with a high BMI (≥ 30kg/m^2^) were significantly at higher risk of being hospitalized.

**Table 1.** Demographic characteristics of hospitalized and non-hospitalized patients for studies reporting mean values.

| Characteristics | Hospitalized |  | Non-hospitalized |  | p-value |
| --- | --- | --- | --- | --- | --- |
|  | Number of studies | Pooled mean/cumulative percent (95% CI) | Number of studies | Pooled mean/cumulative percent (95% CI) |  |
| Male, pooled cumulative per-cent % | 22 | 56.78 (49.43, 64.14) | 21 | 46.16 (38.72, 53.59) | <0.001 |
| Age | 17 | 57.68 (52.20, 63.16) | 18 | 47.93 (42.52, 53.34) | <0.001 |
| BMI | 4 | 29.39 (26.22, 32.56) | 6 | 27.06 (24.01, 30.12) | 0.054 |
| Smoking | 8 | 9.02 (4.76, 13.28) | 7 | 5.83 (1.54, 10.13) | 0.020 |

The missing mean values were replaced with median values, if available.
The pooled results and p-value were obtained by random-effect mixed model with random intercept for study.
Abbreviations: CI, confidence interval

**Figure 1** presents the odds ratio of hospitalized to non-hospitalized COVID-19 positive patients, for several demographic characteristics and comorbidities. Immune deficiency, dementia, coronary artery disease, dyslipidemia, and congestive heart failure were identified as the top comorbidities associated with increased hospitalization risk with odds ratio respectively of 15, 7, 7, 6 and 5. None of the studies reported if the patients with dyslipidemia were stable on their treatment or uncontrolled at the time of the hospitalization. Patients with any renal disease were also at higher risk of being hospitalized, particularly patients with chronic kidney disease. Overall, respiratory conditions increased the hospitalization risk, particularly chronic obstructive pulmonary disease. Adjusted compared to unadjusted analysis showed a statistically significant but lower odds ratio for 3 co-morbidities (chronic kidney disease, hypertension, and diabetes) where the analysis was possible due to sufficient studies available (more than 10).

**Figure 1.**
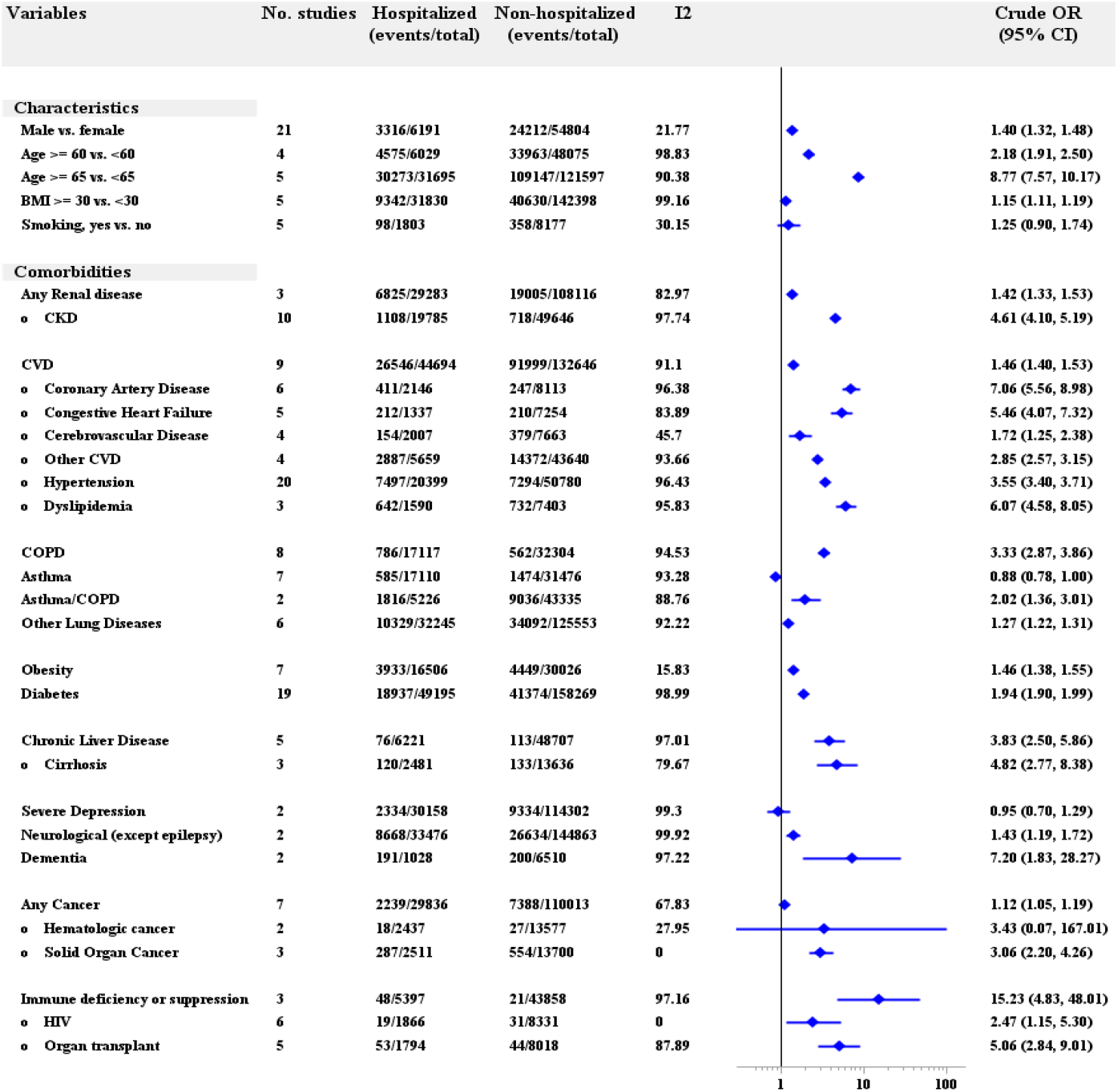
Forest plot for the association of characteristics and some comorbidities with at least 2 studies available and hospitalization.

### 3.2. Mortality risk

Eight studies documented mortality rates and comorbidities [4–6,28–32]. **Table 2** presents the demographics of deceased and living patients. Demographically, male sex was associated with a higher mortality risk and also patients who were older, but these differences didn’t reach statistical significance. Age 60 years and older also increases the odds of death.

**Table 2.** Demographic characteristics of hospitalized and non-hospitalized patients for studies reporting mean values.

| Characteristics | Deceased |  | Living |  | p-value |
| --- | --- | --- | --- | --- | --- |
|  | Number of studies | Pooled mean/cumulative percent (95% CI) | Number of studies | Pooled mean/cumulative percent (95% CI) |  |
| Male | 5 | 58.63 (47.89, 69.38) | 5 | 48.4 (37.65, 59.15) | 0.098 |
| Age | 4 | 72.25 (62.30, 82.20) | 3 | 57.4 (46.56, 68.24) | 0.259 |

The missing mean values were replaced with median values, if available.
The pooled results and p-value were obtained by random-effect mixed model with random intercept for study.
Abbreviations: CI, confidence interval

**Figure 2** presents the odds ratio of deceased and alive COVID-19 positive patients, for several comorbidities. COPD, asthma, CKD, and hypertension were identified as the top comorbidities associated with increased mortality risk among the medical conditions with odds ratios respectively of 25, 12, 8, and 6.

**Figure 2.**
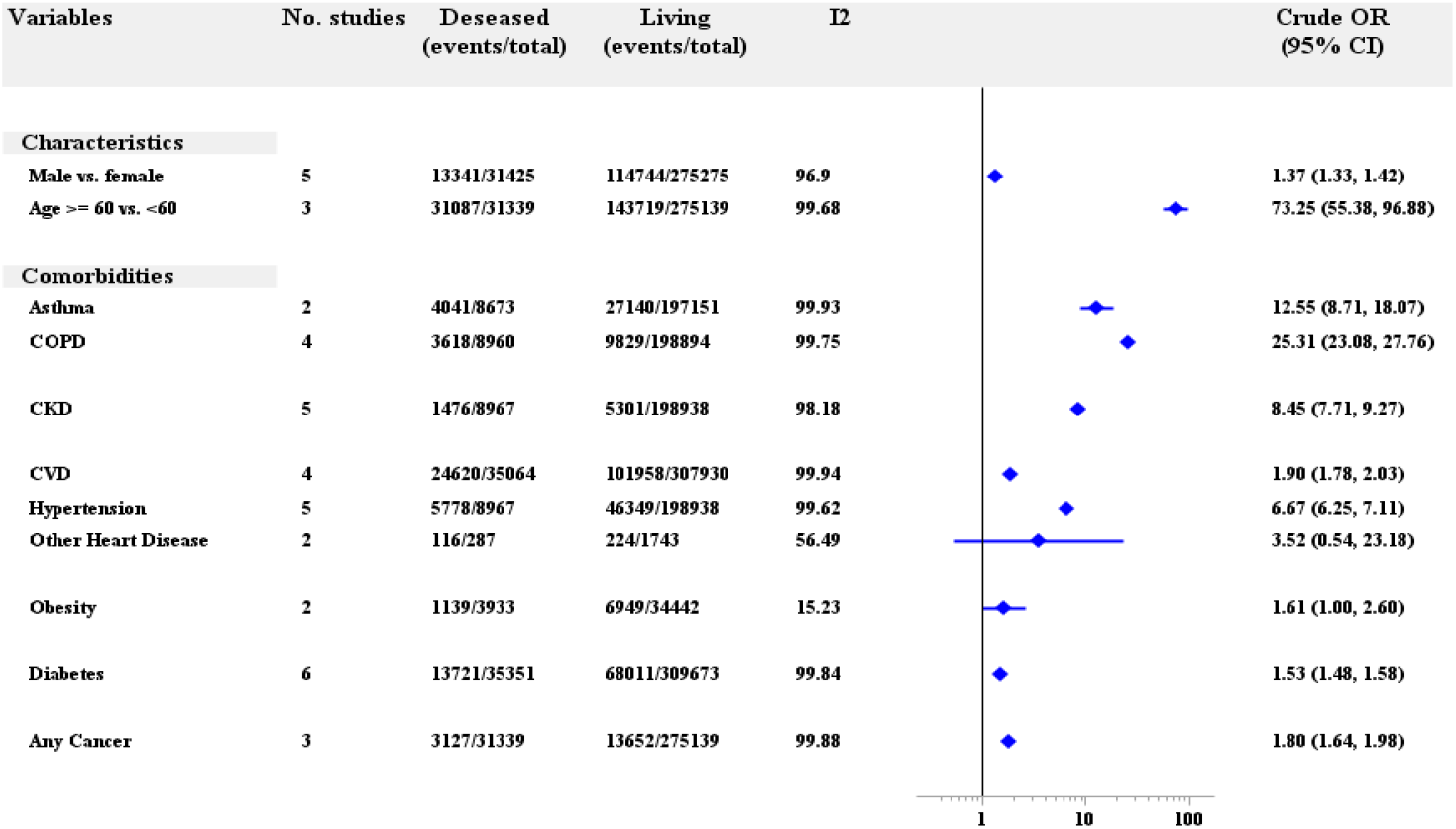
Forest plot for the association of characteristics and some comorbidities with at least 2 studies available and death.

The supplement also presents a radar chart comparing the magnitude of hospitalization and mortality risk for different comorbidities (**Figure S2**).

## 4. Discussion

### Preparing for Pathogen X – Lessons from COVID-19

Pandemics are complex events that involve both political and health aspects [33]. From a health perspective, it is imperative to identify and contain the spread of disease and to care for those who are infected [33]. The COVID-19 pandemic demonstrated that in times of limited healthcare resources it is important for healthcare professionals and healthcare systems to be able to quickly identify individuals or groups who can benefit the most from different interventions. This includes access to public health measures (e.g. masking, disinfectants), therapeutics, vaccines, and treatment options (e.g. ventilation).

Although most pandemic preparedness programs call for increased funding for public health infrastructure, vaccines, therapeutics, and countermeasures, it is crucial to be able to not only have access public health interventions but also to determine:

- Who should receive these interventions?
- How do we prioritize the groups of patients for each intervention?
- How do we evaluate these interventions in the different groups?

To answer these questions for COVID-19 and to prepare for future pandemics our group evaluated the literature to determine if there was clarity on the specific factors that are associated with a higher risk of COVID-19 related outcomes. The goal was to determine if currently published research provided details to address the prioritization of these interventions.

### Incompleteness of risk factor reporting

Through the review of published literature on COVID-19 risk factors a lack of consistency of data reporting was quickly identified. We noted that there was no consistency among all 29 studies in reporting demographics and comorbidities data. For one, some studies had detailed breakdown about the different types of a disease area, while others reported overall proportions creating limitation in the data analysis pooling. This was also the case for BMI and age. We were interested in exploring younger age cut-offs, especially populations such as 50 years and older, however, this data was not available. The majority of studies reported age as a continuous variable, and those that reported age brackets used 60 or 65 years as a cut-off. Second, because of the variation in reporting characteristics, the number of studies used for data analysis was vastly different, sometimes resulting in small sets of data limiting the precision of the estimate and impact statistical significance.

Without more consistency in risk factor reporting for Pathogen X, it would limit the ability to target resources to the groups who are the highest risk and/or could benefit the most from interventions.

### Key gaps in COVID-19 risk factor reporting

#### Lack of population diversity

The representativeness of the results considering that more than half of the studies were conducted in United States and China. Of the 29 studies that were analyzed, 10 were from the United States [5,9– 11,14,19,20,22,30,32], 7 from China [6,8,15,16,23,25,26] and only 12 from a variety of other countries including, Canada, Denmark, England, Gabon, Indonesia, Japan, Korea, Malaysia, Mexico, Poland, and Scotland [4,7,12,13,17,18,21,24,28,29,31].

#### Lack of variant publication

There were different periods of study publications: 17 studies were published in 2021 [4,5,7,9,12– 14,18,20,21,25,27–32] and 12 in 2020 [6,8,10,11,15–17,22–24,26]. It was not possible to associate a relevant COVID-19 variant with each study. As a pandemic evolves, the priority structure for resources will likely change. Without variant data, it is impossible to determine if future variants pose a lower, similar or higher risk to specific patient populations.

#### Lack of vaccination data

It is important to discuss the challenges in trying to analyze the effect of vaccines, as only 17 studies were published in 2021 and from those selected for full read and data collection, it was not described whether vaccines were available (and if so, which type of vaccines) at the time of the outcome measurement being reported. With an evolving pandemic the number of vaccines given, the antigen dosing, spacing and breakthrough infections can provide further guidance on where to allocate resources.

The COVID-19 pandemic data has demonstrated the need for a standardized and comprehensive risk factor reporting structure to allow researchers, healthcare professionals and healthcare systems for preparing for Pathogen X.

### It is time for CLARITY

To address the underreporting of key data for the evaluation of risk factors, we proposed a comprehensive structure for risk factor reporting for studies for pandemic pathogens. The CLARITY model can help to address key gaps that were identified when evaluating the COVID-19-related risk factor published literature. **Figure 3** reviews the components of this model.

**Figure 3.**
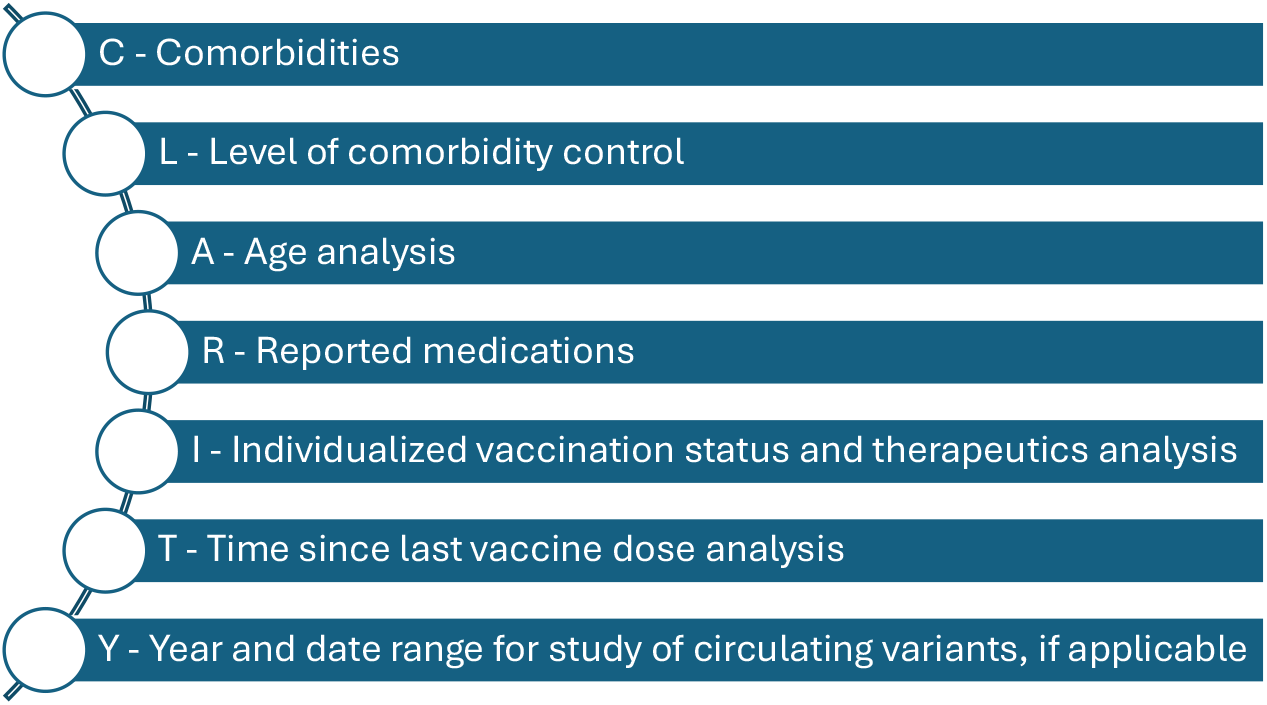
The CLARITY model for reporting of pandemic-related risk factors.

#### Comorbidities

The first component of the model is a comprehensive reporting of the different comorbidities of the patient population. Many of the published studies were designed to evaluate the impact of one comorbidity on COVID-19 outcome risk but did not provide data if the patient population had other comorbidities that further increased the risk. It was impossible to determine the level of ‘comorbidity stacking’ on the person’s risk of severe COVID-19. Most of the reported data was able to identify if a comorbidity (e.g. COPD) increased their risk of severe COVID-19 but could not quantify the potential increased risk level if the individual had additional risk factors (e.g. cardiovascular disease, obesity, hypertension, type 2 diabetes). This is a significant gap as others have reported an increase in the number of comorbidities is associated with a higher risk of COVID-19 related death, invasive mechanical ventilation, and death [34].

A comprehensive list of comorbidities for the evaluated population for Pathogen X would help to identify the extent that each contribute to the individual’s risk of severe outcomes.

#### Level of comorbidity control

Currently published research on comorbidity risk factors for COVID-19 outcomes, did not consistently report the level of disease control. Although it was possible to determine if the presence of a comorbidity increased the risk of severe COVID-19 related outcomes, we could not quantify if the risk was higher in a person had poorer comorbidity control. For example, much of the published research would indicate that chronic kidney disease (CKD) increases the risk of severe COVID-19, but measures such as glomerular filtration rate or albumin : creatinine ratio were not constantly reported.

For Pathogen X, it would be important to not only report the presence of a comorbidity but the overall level of disease control. This could allow further targeting of pandemic resources to the highest risk group with each comorbidity.

#### Age analysis

The impact of age on severe outcomes from infectious disease is seen across multiple pathogens. Different age groups infected with influenza, RSV, pneumococcus, and COVID-19 have variable risk of severe outcomes from these infections. Many of the published research on COVID-19 related risk factors did not provide detailed analysis of the age group of affected individuals. Studies commonly reported age as a continuous variable, and others used specific age cut-offs that limited further analysis.

Age is likely to impact the overall risk of severe outcomes from Pathogen X. Detailed report of age (mean, median, standard deviation) can allow for further risk calculation.

#### Reported medications

There was a lack of consistent reporting on the medications taken by individuals with severe COVID-19 in the published research. This makes it impossible to further stratify both risk and possible benefits based on the specific medications or combination of medications.

Providing a full analysis of each of the medications taken by individuals infected with Pathogen X, would allow for further risk/benefit stratification.

#### Individualized vaccination status and therapeutics analysis

A key gap in the published research is the lack of data on the vaccination status of individuals at risk of severe COVID-19 outcomes. The reviewed studies on COVID-19 risk factors did not allow for any additional analysis on the impact of vaccination, specific vaccine, and number of doses on COVID-19 related risk in individuals with comorbidities. With some individuals being given therapeutics such as monoclonal antibodies, it would be helpful to determine their impact on overall risk. With limited healthcare resources, it would be useful to determine if vaccination or therapeutic lowers the risk so significantly in some groups that it could lead to reallocation of these resources.

Capturing additional vaccination information for each individual, such as flu, pneumococcal, RSV, BCG, etc. in the case of COVID-19, could equally allow for analysis of impact of vaccination on disease severity risk. Vaccination details and therapeutic information could help to guide healthcare professionals and health systems to further classify risk for Pathogen X.

#### Time since last vaccine dose analysis

The greater the time since the last vaccine dose is associated with a lower overall vaccine effectiveness [35]. It is not simply the vaccine type, dose and the number of vaccine doses, but also the time since the last vaccine dose that can significantly impact the overall risk.

If multiple vaccine doses are required to reduce the risk of Pathogen X, time since last vaccine dose will provide crucial analysis for those with severe outcomes.

#### Year and date range for study of circulating variants, if applicable

The continued evolution of the SARS-CoV-2 virus, there has been differences in the risk of severe COVID-19 outcomes. Without information on the time of collected data, it would be impossible to determine the impact of circulating variants.

If Pathogen X develops variants with different levels of infectivity and outcome risk, it will be crucial to have date analysis to determine the impact of variants on overall risk.

## 5. Conclusion

Our review emphasizes the urgent need for a coordinated global effort to enhance pandemic preparedness, particularly for future threats posed by unknown pathogens, or Disease X. The COVID-19 pandemic has revealed critical gaps and inconsistencies in the reporting of risk factors, which impede effective resource allocation and response strategies. Our findings highlight the necessity of standardizing reported data on comorbidities, age and BMI brackets, disease control levels, medication use, vaccination status, and the impact of viral variants.

The proposed CLARITY model addresses these deficiencies by providing a comprehensive framework for risk factor reporting, which includes detailed comorbidity profiles, disease control metrics, age analyses, medication and vaccination data, and variant-specific information. Implementing this model will enable healthcare professionals, researchers, public health officials and healthcare systems to more accurately identify and prioritize high-risk populations, optimize resource distribution, and improve outcomes during future pandemics.

Furthermore, the review underscores the importance of understanding public attitudes, knowledge, and behaviours regarding vaccines, as these factors significantly influence the effectiveness of vaccination campaigns and public health interventions. By adopting the CLARITY model, researchers, healthcare professionals, and policymakers can develop more informed and targeted strategies to mitigate the impact of future pandemics. Ultimately, this will contribute to building more resilient healthcare systems capable of responding swiftly and effectively to emerging infectious threats.

## Data Availability

All data produced in the present study are available upon reasonable request to the authors.

## Acknowledgments

We would like to acknowledge the assistance of Dr. Claire Harris, MD FRCPC (University of British Columbia) with the initial literary search review, and the non-profit society RESPIPLUS for providing a key role in planning committee development, logistics support and engaging key stakeholders regarding the results of this study.

