## Supplement for "Preparing for Disease X requires CLARITY – Lessons learned from COVID-19 – A systematic Review"

### 1. Additional Information on Methodology

#### Literature search strategy

A systematic literature search was conducted using Ovid MEDLINE(R) and Epub databases to identify manuscripts specific to COVID-19 and high-risk populations, published between December 2019 and March 2023. The key words "coronavirus", "COVID-19", "vulnerable populations", "high-risk", "death", "ICU" and "comorbid" were used in various combinations. Manuscripts ahead of print, in-process, in-data-review, and other non-indexed citations were all searched and collected if deemed eligible. Articles relevant to other members of the coronavirus family such as Severe Acute Respiratory Syndrome Coronavirus 2 (SARS-CoV-2) and Middle East respiratory syndrome (MERS) were excluded from the final list of selected articles for screening, which included 440 eligible articles. A PRISMA Checklist is available in **Table S1** and **S2**, and the literature search strategy is summarized in **Table S3** of this Supplement.

#### Study eligibility and selection

The inclusion criteria were observational studies, including retrospective and prospective studies, describing the association between COVID-19 or SARS-COV-2 and comorbid conditions, and reporting outcomes such as hospitalization and mortality; only articles published in English; predominantly in adult populations (less than 10% pediatric population). Exclusion criteria were meta-analysis studies; interventional studies such as clinical trials and drug treatments; studies investigating prognostic factors, sex differences, socioeconomic factors, pathogenesis, and mechanisms of action; impact of COVID-19 on disease severity; and overall well-being. Three reviewers working independently screened records using Covidence systematic review software, Veritas Health Innovation, available at www.covidence.org. Selected studies were assessed for risk of bias using the Cochrane Risk Of Bias due to Missing Evidence ROB-ME tool (© 2023 under Creative Commons License).

#### Data extraction

The data was extracted by one reviewer and included study type, number of patients, age, body mass index (BMI), sex, smoking, comorbidities, mortality, hospitalization, ICU admission, oxygen therapy, and intubation. The age extracted from the studies were grouped based on 2 cut-offs: those who were 60 years and older or 65 years and older, based on the reported ages per study. BMI was grouped into 2 categories: less than 30 kg/m$^2$ or greater than or equal to 30 kg/m$^2$.

#### Statistical analysis

Participant characteristics were summarized using pooled mean or cumulative percent with 95% confidence intervals (CIs), as appropriate. Some studies reported mean values, some reported median, and the missing mean values were replaced with median values, if available.

We used a random-effects (random intercept for study) generalized logistic mixed effects model (PROC GLIMMIX in SAS) to estimate crude odds ratios (ORs) and 95% CI of hospitalization (versus non-hospitalization) or death (versus living) associated with participant characteristics and comorbidities. For characteristics, estimates were calculated for the studies reported sex, smoking status, and the category continuous characteristics variables (age and/or BMI). In the analysis of comorbidities, we selected the comorbidities reported in at least 2 studies. Adjusted ORs (95% CIs) were estimated using multiple GLIMMIX model for the comorbidities with at least 10 studies available, adjusted for sex and mean age. The available numbers of studies in the multiple analysis were different from those in the univariate analysis because some studies were not reported mean age but reported category age, and these studies were excluded in the multiple analysis. To test for heterogeneity of ORs across studies, we used Breslow-

Day Test to calculate the $I^2$ statistic. Radar chart was created to compare the magnitude of association of different comorbidities for both hospitalization and death.
Statistical significance was defined as two-sided p-value $< 0.05$. Statistical analyses were conducted using the SAS version 9.4 software (TS1M5) (SAS Institute Inc., Cary, NC, USA, 2016).

<u>**Supplementary Tables and Figures**</u>

**Table S1: PRISMA 2020 Checklist**

**Table S2: PRISMA 2020 Abstract Checklist**

**Table S3: Literary Search Strategy**

**Figure S1: PRISMA Flowchart for Study Selection**

**Table S4: Characteristics of Included Studies**

**Figure S2. Radar chart to compare the magnitude of association of different comorbidities for both hospitalization and death**

**Supplement Table S1.** PRISMA_2020_Checklist

| Section and Topic | Item # | Checklist item | Location where item is reported |
|---|---|---|---|
| **TITLE** | | | |
| Title | 1 | Identify the report as a systematic review. | Page no 1 |
| **ABSTRACT** | | | |
| Abstract | 2 | See the PRISMA 2020 for Abstracts checklist. | Table S2 |
| **INTRODUCTION** | | | |
| Rationale | 3 | Describe the rationale for the review in the context of existing knowledge. | Page no 2 |
| Objectives | 4 | Provide an explicit statement of the objective(s) or question(s) the review addresses. | Page no 2 |
| **METHODS** | | | |
| Eligibility criteria | 5 | Specify the inclusion and exclusion criteria for the review and how studies were grouped for the syntheses. | Table S3 |
| Information sources | 6 | Specify all databases, registers, websites, organisations, reference lists and other sources searched or consulted to identify studies. Specify the date when each source was last searched or consulted. | Table S3 |
| Search strategy | 7 | Present the full search strategies for all databases, registers and websites, including any filters and limits used. | Table S3 |
| Selection process | 8 | Specify the methods used to decide whether a study met the inclusion criteria of the review, including how many reviewers screened each record and each report retrieved, whether they worked independently, and if applicable, details of automation tools used in the process. | Supplementary Material |
| Data collection process | 9 | Specify the methods used to collect data from reports, including how many reviewers collected data from each report, whether they worked independently, any processes for obtaining or confirming data from study investigators, and if applicable, details of automation tools used in the process. | Supplementary Material |
| Data items | 10a | List and define all outcomes for which data were sought. Specify whether all results that were compatible with each outcome domain in each study were sought (e.g. for all measures, time points, analyses), and if not, the methods used to decide which results to collect. | Supplementary Material |

| Section and Topic | Item # | Checklist item | Location where item is reported |
|---|---|---|---|
| | 10b | List and define all other variables for which data were sought (e.g. participant and intervention characteristics, funding sources). Describe any assumptions made about any missing or unclear information. | Not aplicable |
| Study risk of bias assessment | 11 | Specify the methods used to assess risk of bias in the included studies, including details of the tool(s) used, how many reviewers assessed each study and whether they worked independently, and if applicable, details of automation tools used in the process. | Supplementary Material |
| Effect measures | 12 | Specify for each outcome the effect measure(s) (e.g. risk ratio, mean difference) used in the synthesis or presentation of results. | Supplementary Material |
| Synthesis methods | 13a | Describe the processes used to decide which studies were eligible for each synthesis (e.g. tabulating the study intervention characteristics and comparing against the planned groups for each synthesis (item #5)). | Supplementary Material |
| | 13b | Describe any methods required to prepare the data for presentation or synthesis, such as handling of missing summary statistics, or data conversions. | Not applicable |
| | 13c | Describe any methods used to tabulate or visually display results of individual studies and syntheses. | Supplementary Material |
| | 13d | Describe any methods used to synthesize results and provide a rationale for the choice(s). If meta analysis was performed, describe the model(s), method(s) to identify the presence and extent of statistical heterogeneity, and software package(s) used. | Supplementary Material |
| | 13e | Describe any methods used to explore possible causes of heterogeneity among study results (e.g. subgroup analysis, meta-regression). | Supplementary Material |
| | 13f | Describe any sensitivity analyses conducted to assess robustness of the synthesized results. | Supplementary Material |
| Reporting bias assessment | 14 | Describe any methods used to assess risk of bias due to missing results in a synthesis (arising from reporting biases). | Supplementary Material |
| Certainty assessment | 15 | Describe any methods used to assess certainty (or confidence) in the body of evidence for an outcome. | Supplementary Material |
| **RESULTS** | | | |
| Study | 16a | Describe the results of the search and selection process, from the number of records identified in the search to the | Page no 2, |

| Section and Topic | Item # | Checklist item | Location where item is reported |
|---|---|---|---|
| selection | | number of studies included in the review, ideally using a flow diagram. | Figure S1 |
| | 16b | Cite studies that might appear to meet the inclusion criteria, but which were excluded, and explain why they were excluded. | Page no 2 |
| Study characteristics | 17 | Cite each included study and present its characteristics. | Page no 2<br>Table S4 |
| Risk of bias in studies | 18 | Present assessments of risk of bias for each included study. | Page 2 |
| Results of individual studies | 19 | For all outcomes, present, for each study (a) summary statistics for each group (where appropriate) and (b) an effect estimates and its precision (e.g. confidence/credible interval), ideally using structured tables or plots. | Tables 1, 2, Figures 1-2 |
| Results of syntheses | 20a | For each synthesis, briefly summarise the characteristics and risk of bias among contributing studies. | Page 3 |
| | 20b | Present results of all statistical syntheses conducted. If meta-analysis was done, present for each the summary estimate and its precision (e.g. confidence/credible interval) and measures of statistical heterogeneity. If comparing groups, describe the direction of the effect. | Figures 1-2 |
| | 20c | Present results of all investigations of possible causes of heterogeneity among study results. | Page 4 |
| | 20d | Present results of all sensitivity analyses conducted to assess the robustness of the synthesized results. | Page 4 |
| Reporting biases | 21 | Present assessments of risk of bias due to missing results (arising from reporting biases) for each synthesis assessed. | Page 4 |
| Certainty of evidence | 22 | Present assessments of certainty (or confidence) in the body of evidence for each outcome assessed. | Tables 1, 2 Figures 1-2 |
| **DISCUSSION** | | | |
| Discussion | 23a | Provide a general interpretation of the results in the context of other evidence. | Page no 5-6 |
| | 23b | Discuss any limitations of the evidence included in the review. | Page no 5-6 |
| | 23c | Discuss any limitations of the review processes used. | Page no 5-6 |
| | 23d | Discuss implications of the results for practice, policy, and future research. | Page no 6-9 |

| Section and Topic | Item # | Checklist item | Location where item is reported |
|---|---|---|---|
| **OTHER INFORMATION** | | | |
| Registration and protocol | 24a | Provide registration information for the review, including register name and registration number, or state that the review was not registered. | Page no 2 |
| | 24b | Indicate where the review protocol can be accessed, or state that a protocol was not prepared. | Supplementary Material |
| | 24c | Describe and explain any amendments to information provided at registration or in the protocol. | Not applicable |
| Support | 25 | Describe sources of financial or non- financial support for the review, and the role of the funders or sponsors in the review. | Page no 9 |
| Competing interests | 26 | Declare any competing interests of review authors. | Title page |
| Availability of data, code and other materials | 27 | Report which of the following are publicly available and where they can be found template data collection forms; data extracted from included studies; data used for all analyses; analytic code; any other materials used in the review. | Not applicable |

*From* Page MJ, McKenzie JE, Bossuyt PM, Boutron I, Hoffmann TC, Mulrow CD, et al. The PRISMA 2020 statement an updated guideline for reporting systematic reviews. BMJ 2021;372n71. doi 10.1136/bmj.n71 For more information, visit http//www.prisma.statement.org/

**Supplementary Table S2.** PRISMA 2020 Abstract Checklist

| Section and Topic | Item # | Checklist item | Reported (Yes/No) |
|---|---|---|---|
| **TITLE** | | | |
| Title | 1 | Identify the report as a systematic review. | yes |
| **BACKGROUND** | | | |
| Objectives | 2 | Provide an explicit statement of the main objective(s) or question(s) the review addresses. | yes |
| **METHODS** | | | |
| Eligibility criteria | 3 | Specify the inclusion and exclusion criteria for the review. | yes |
| Information sources | 4 | Specify the information sources (e.g. databases, registers) used to identify studies and the date when each was last searched. | yes |
| Risk of bias | 5 | Specify the methods used to assess risk of bias in the included studies. | no |
| Synthesis of results | 6 | Specify the methods used to present and synthesise results. | no |
| **RESULTS** | | | |
| Included studies | 7 | Give the total number of included studies and participants and summarise relevant characteristics of studies. | yes |
| Synthesis of results | 8 | Present results for main outcomes, preferably indicating the number of included studies and participants for each. If meta-analysis was done, report the summary estimate and confidence/credible interval. If comparing groups, indicate the direction of the effect (i.e. which group is favoured). | yes |
| **DISCUSSION** | | | |
| Limitations of evidence | 9 | Provide a brief summary of the limitations of the evidence included in the review (e.g. study risk of bias, inconsistency and imprecision). | yes |
| Interpretation | 10 | Provide a general interpretation of the results and important implications. | yes |
| **OTHER** | | | |
| Funding | 11 | Specify the primary source of funding for the review. | yes |
| Registration | 12 | Provide the register name and registration number. | yes |

**Supplementary Table S3. Literature Search Strategy**

| # | Query | Initial search on June 13, 2022 |
|---|---|---|
| 1 | exp Coronavirus/ | 141,271 |
| 2 | exp Coronavirus Infections/ | 177,528 |
| 3 | (coronavirus* or corona virus* or OC43 or NL63 or 229E or HKU1 or HCoV* or ncov* or covid* or sars-cov* or sarscov* or Sars-coronavirus* or Severe Acute Respiratory Syndrome Coronavirus* or "Kawasaki like paediatric inflammatory multisystem syndrome" or "Kawasaki like pediatric inflammatory multisystem syndrome" or "PIMS-TS" or "Kawa-COVID-19" or "MIS-C" or "multisystem inflammatory syndrome in children" or pediatric multisystem inflammatory disease).mp. | 281,486 |
| 4 | (or/1-3) and ((20191* or 202*).dp. or 20190101:20301231.(ep).) [this set is the sensitive/broad part of the search] | 268,543 |
| 5 | 4 not (SARS or SARS-CoV or MERS or MERS-CoV or Middle East respiratory syndrome or camel* or dromedar* or equine or coronary or coronal or covidence* or covidien or influenza virus or HIV or bovine or calves or TGEV or feline or porcine or BCoV or PED or PEDV or PDCoV or FIPV or FCoV or SADS-CoV or canine or CCov or zoonotic or avian influenza or H1N1 or H5N1 or H5N6 or IBV or murine corona*).mp. [line 5 removes SARS, MERS, and veterinary noise from the sensitive/broad search results] | 96,501 |
| 6 | ((pneumonia or covid* or coronavirus* or corona virus* or ncov* or 2019-ncov or sars*).mp. or exp pneumonia/) and Wuhan.mp. [Early articles about the outbreak] | 6,992 |
| 7 | (2019-ncov or ncov19 or ncov-19 or 2019-novel CoV or sars-cov2 or sars-cov-2 or sarscov2 or sarscov-2 or SARS-2-nCoV or SARS-2-Cov or SARS-COV-19 or Sars-coronavirus2 or Sars-coronavirus-2 or SARS 2 coronavirus* or Severe Acute Respiratory Syndrome-CoV-2 or SARS-like coronavirus* or coronavirus-19 or covid19 or covid-19 or covid 2019 or ((novel or new or nouveau) adj2 (CoV or nCoV or covid or coronavirus* or corona virus or Pandemi*2)) or ((covid or covid19 or covid-19 or SARS-CoV-2) and pandemic*2) or (coronavirus* and pneumonia)).mp. [specific to Covid-19, Covid pneumonia, Covid pandemic] | 264,757 |
| 8 | (2019-ncov or ncov19 or ncov-19 or 2019-novel CoV or sars-cov2 or sars-cov-2 or sarscov2 or sarscov-2 or SARS-2-nCoV or SARS-2-Cov or SARS-COV-19 or Sars-coronavirus2 or Sars-coronavirus-2 or SARS 2 coronavirus* or Severe Acute Respiratory Syndrome-CoV-2 or SARS-like coronavirus* or coronavirus-19 or covid19 or covid-19 or covid 2019 or ((novel or new or nouveau) adj2 (CoV or nCoV or covid or coronavirus* or corona virus or Pandemi*2)) or ((covid or covid19 or covid-19 or SARS-CoV-2) and pandemic*2) or (coronavirus* and pneumonia)).mp. [specific to Covid-19, Covid pneumonia, Covid pandemic] | 264,757 |

| | | |
|---|---|---|
| 9 | (COVID-19 or SARS-CoV-2).rx,px,ox,rn. or (COVID-19 or COVID-19 serotherapy or ORF7b protein, SARS-CoV-2 or ORF6 protein, SARS-CoV-2 or ORF8 protein, SARS-CoV-2 or pediatric multisystem inflammatory disease, COVID-19 related or envelope protein, SARS-CoV-2 or ORF7a protein, SARS-CoV-2 or spike protein, SARS-CoV-2 or ORF3a protein, SARS-CoV-2 or COVID-19 drug treatment or severe acute respiratory syndrome coronavirus 2 or membrane protein, SARS-CoV-2 or ORF1ab polyprotein, SARS-CoV-2 or nucleocapsid protein, Coronavirus or COVID-19 vaccine or COVID-19 diagnostic testing).os,ps,rn,rs. [Relevant Suppl. Concepts listed in MeSH 2021 Browser] | 24,008 |
| 10 | or/5-9 [Lines 5 to 9 are specific/relevant to the Covid-19 outbreak] | 268,409 |
| 11 | 10 and 20191201:20301231.(dt). [only include records created for PubMed/MEDLINE from 1 December 2019 onwards] | 266,254 |
| 12 | *Vulnerable Populations/ or *Severity of Illness Index/ or exp *Risk Factors/ or ((exp *Risk/ or exp *Risk Factors/) and (*Hospital Mortality/ or exp *Death/ or exp *Intensive Care Units/)) | 31,174 |
| 13 | ((patient* or population* or group* or comorbid*) adj3 ((vulnerable or high-risk or "high risk") adj2 (disease sever* or death* or ICU or intensive care or mortality))).ti,kf. or ((patient* or population* or group* or comorbid*) adj3 ((vulnerable or high-risk or "high risk") adj3 (disease sever* or death* or ICU or intensive care or mortality))).ab. /freq=2 | 316 |
| 14 | 12 or 13 | 31,477 |
| 15 | 11 and 14 | 848 |
| 16 | exp Epidemiologic Studies/ | 2,963,616 |
| 17 | (((case or cases) and (control or controls)) or cohort or "follow up study" or "follow-up study" or "observational study" or longitudinal or retrospective or "cross sectional" or compared or multivariate or epidemiologic study).tw,kf. | 5,899,692 |
| 18 | 16 or 17 | 7,039,324 |
| 19 | 15 and 18 | 437 |
| **20** | **19 and 20220610:20230323.(dt). (Updated query to March 13, 2023)** | **3** |

**Supplementary Figure S1. PRISMA flowchart for study selection**

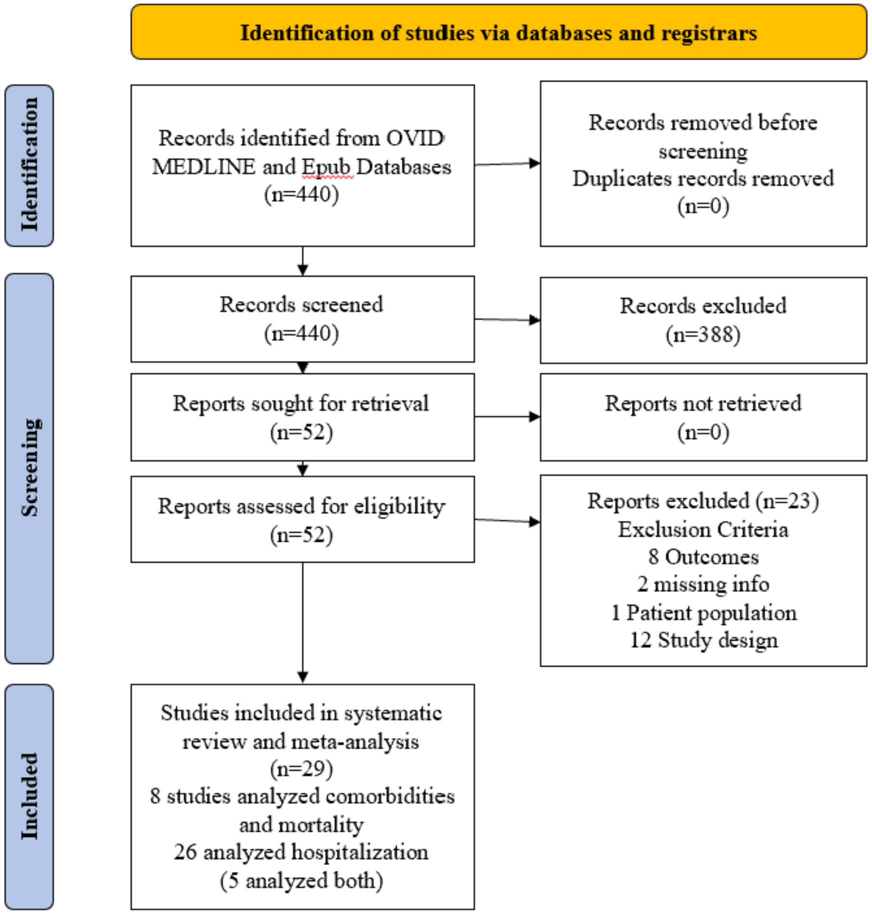

**Supplementary Table S4. Characteristics of Included Studies**

| Study Year | Source [Reference] | Study Name | Country | Study design | Total Number |
|---|---|---|---|---|---|
| 2021 | Ge et al [2] | Association of pre-existing comorbidities with mortality and disease severity among 167,500 individuals with COVID-19 in Canada | Canada | Population-based case-control study | 167,500 |
| 2021 | Ip et al [3] | Atrial Fibrillation as a Predictor of Mortality in High Risk COVID-19 Patients | United States | Multicenter cohort study | 171 |
| 2021 | Hasani et al [10] | Comorbidities and clinical features related to severe outcomes among COVID-19 cases in Selangor, Malaysia | Malaysia | Retrospective case-control study | 1,287 |
| 2020 | McKeigue et al [22] | Rapid Epidemiological Analysis of Comorbidities and Treatments as risk factors for COVID-19 in Scotland (REACT-SCOT) | Scotland | Population-based case-control study | 41,220 |
| 2021 | Pandita et al [25] | Predictors of severity and mortality among patients hospitalized with COVID-19 in Rhode Island | United States | Retrospective Cohort Study | 259 |
| 2020 | Kalligeros et al [8] | Association of Obesity with Disease Severity Among Patients with Coronavirus Disease 2019 | United States | Retrospective Cohort Study | 103 |
| 2021 | Ando et al [18] | Impact of overlapping risks of type 2 diabetes and obesity on coronavirus disease severity in the United States | United States | Cohort Study | 28,093 |
| 2021 | Gao et al [26] | Associations between body-mass index and COVID-19 severity in 6·9 million people in England: a prospective, community-based, cohort study | England | Prospective Cohort Study | 6,910,695 |
| 2020 | Ji et al [15] | Effect of Underlying Comorbidities on the Infection and Severity of COVID-19 in Korea: a Nationwide Case-Control Study | Korea | Retrospective case-control Study | 7,341 |
| 2020 | Zhang et al [24] | Risk Factors for Poor Outcomes of Diabetes Patients With COVID-19: A Single-Center, Retrospective Study in Early Outbreak in China | China | Single Center Retrospective Cohort Study | 52 |

| | | | | | |
|---|---|---|---|---|---|
| 2020 | Deng et al [21] | Obesity as a Potential Predictor of Disease Severity in Young COVID-19 Patients: A Retrospective Study | China | Retrospective Cohort Study | 65 |
| 2021 | Stachura et al [5] | A clinical profile and factors associated with severity of the disease among Polish patients hospitalized due to COVID-19 — an observational study | Poland | An observational study | 100 |
| 2021 | Martos-Benítez et al [27] | Chronic comorbidities and clinical outcomes in patients with and without COVID-19: a large population-based study using national administrative healthcare open data of Mexico | Mexico | Retrospective case-control Study | 38,324 |
| 2020 | Gottlieb et al [9] | Clinical Course and Factors Associated With Hospitalization and Critical Illness Among COVID-19 Patients in Chicago, Illinois | United Sates | Retrospective, registry-based cohort study | 8,673 |
| 2020 | Bhargava et al [20] | Predictors for Severe COVID-19 Infection | United States | Retrospective observational study | 197 |
| 2021 | Hirashima et al [16] | Factors significantly associated with COVID-19 severity in symptomatic patients: A retrospective single-center study | Japan | Retrospective Cohort Study | 61 |
| 2021 | Garibaldi et al [28] | Patient Trajectories Among Persons Hospitalized for COVID-19_A Cohort Study | United States | Retrospective Cohort Study | 832 |
| 2021 | Fernandes et al [7] | Adult COVID-19 Patients Cared for in a Pediatric ICU Embedded in a Regional Biothreat Center: Disease Severity and Outcomes | United States | Retrospective cohort study | 37 |
| 2020 | Chen et al [4] | Risk factors for death in 1859 subjects with COVID-19 | China | Cohort Study | 1,859 |
| 2021 | Gunadi et al [29] | Association between prognostic factors and the outcomes of patients infected with SARS-CoV-2 harboring multiple spike protein mutations | Indonesia | Retrospective Cohort Study | 51 |
| 2020 | Yao et al [6] | A retrospective study of risk factors for severe acute respiratory syndrome coronavirus 2 infections in hospitalized adult patients | China | Retrospective Cohort Study | 108 |

| | | | | | |
|---|---|---|---|---|---|
| 2020 | Axelrad et al [17] | From the American Epicenter: Coronavirus Disease 2019 in Patients with Inflammatory Bowel Disease in the New York City Metropolitan Area | United Sates | Cohort Study | 83 |
| 2020 | Zhang et al [14] | Do underlying cardiovascular diseases have any impact on hospitalised patients with COVID-19? | China | Retrospective Cohort Study | 541 |
| 2021 | Iroungou et al [11] | Demographic and Clinical Characteristics Associated With Severity, Clinical Outcomes, and Mortality of COVID-19 Infection in Gabon | Gabon | Retrospective Cross-sectional Study | 313 |
| 2020 | Hu et al [13] | Early prediction and identification for severe patients during the pandemic of COVID-19: A severe COVID-19 risk model constructed by multivariate logistic regression analysis | China | Retrospective non-interventional study, | 40 |
| 2021 | Lokken et al [12] | Disease severity, pregnancy outcomes, and maternal deaths among pregnant patients with severe acute respiratory syndrome coronavirus 2 infection in Washington State | United Sates | Multicenter retrospective cohort study | 240 |
| 2021 | Mehta et al [30] | Risk Factors Associated With SARS-CoV-2 Infections, Hospitalization, and Mortality Among US Nursing Home Residents | United States | Cohort Study | 137,119 |
| 2021 | Cordtz et al [19] | Incidence and severeness of COVID-19 hospitalisation in patients with inflammatory rheumatic disease: a nationwide cohort study from Denmark | Denmark | Cohort Study | 47 |
| 2021 | Geng et al [23] | Risk factors for developing severe COVID-19 in China: an analysis of disease surveillance data | China | Cross-sectional study | 12,647 |

**Supplementary Figure S2. Radar chart to compare the magnitude of association of different comorbidities for both hospitalization and death**

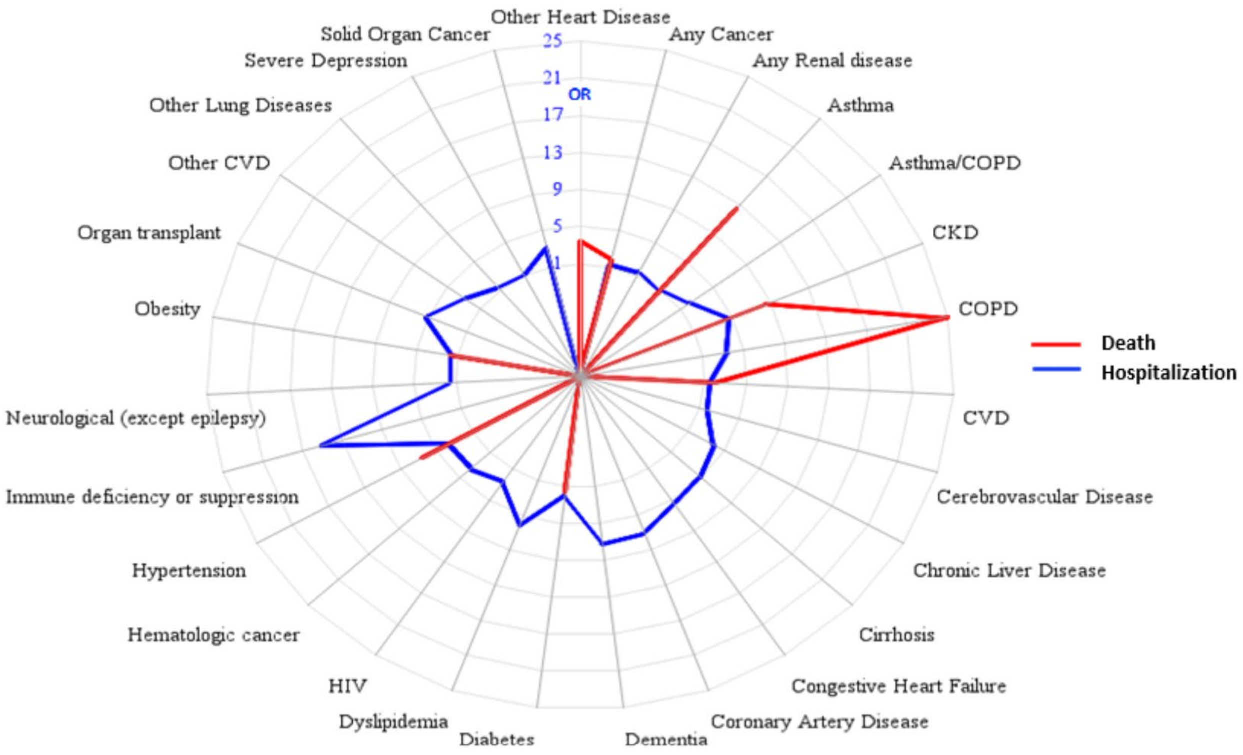

Abbreviations: CKD, Chronic Kidney Disease; COPD, Chronic Obstructive Pulmonary Disease; CVD, Cardiovascular Disease; HIV, human immunodeficiency virus
